# Oral insulin immunotherapy in children at risk for type 1 diabetes in a randomized trial

**DOI:** 10.1101/2020.06.12.20129189

**Authors:** Robin Assfalg, Jan Knoop, Kristi L. Hoffman, Markus Pfirrmann, Jose Maria Zapardiel-Gonzalo, Anna Hofelich, Anne Eugster, Marc Weigelt, Claudia Matzke, Julia Reinhardt, Yannick Fuchs, Melanie Bunk, Andreas Weiss, Markus Hippich, Kathrin Halfter, Stefanie M. Hauck, Jörg Hasford, Joseph F. Petrosino, Peter Achenbach, Ezio Bonifacio, Anette-Gabriele Ziegler

## Abstract

**Background:** Oral administration of antigen can induce immunological tolerance. Insulin is a key autoantigen in childhood type 1 diabetes with insulin autoimmunity often appearing in the first years of life. Here, oral insulin was given as antigen-specific immunotherapy before the onset of autoimmunity in children from age 6 months to assess its safety and actions on immunity and the gut microbiome.

**Methods:** A phase I/II randomized controlled trial was performed in 44 islet autoantibody-negative children aged 6 months to 2 years with genetic risk for type 1 diabetes. Children were randomized 1:1 to daily oral insulin (7.5 mg with dose escalation to 67.5 mg) or placebo for 12 months. Primary outcome was safety and immune efficacy pre-specified as hypoglycemia and induction of antibody or T cell responses to insulin, respectively.

**Results:** Oral insulin was well tolerated with no changes in metabolic variables. Immune responses to insulin were observed in both children who received insulin (55%) and placebo (67%), and were modified by the *INSULIN* gene. Among children with type 1 diabetes-susceptible *INSULIN* genotype, antibody responses to insulin were more frequent in insulin-treated (cumulative response, 75.8%) as compared to placebo-treated children (18.2%; *P*=0.0085), and T cell responses to insulin were modified by treatment-independent inflammatory episodes. Changes in the microbiome were related to *INSULIN* genotype.

**Conclusion:** The study demonstrated that oral insulin immunotherapy in young genetically at-risk children was safe and engaged the adaptive immune system in an *INSULIN* genotype-dependent manner, and linked inflammatory episodes to the activation of insulin-responsive T cells.

**Trial registration:** Clinicaltrials.gov NCT02547519

**Funding:** German Center for Diabetes Research (DZD e.V.), Juvenile Diabetes Research Foundation (JDRF, grant 1-SRA-2018-546-S-B), Federal Ministry of Education and Research (BMBF, grant FKZ01KX1818).

## Introduction

Type 1 diabetes results from an autoimmune destruction of insulin-producing β cells in the pancreatic islets of Langerhans, and is characterized by circulating islet autoantibodies to β cell antigens (1, 2). Insulin is a key early autoantigen in childhood diabetes (3, 4). Autoimmunity against insulin often appears in genetically susceptible children aged 9 months–3 years, with a peak incidence at 9–12 months of age (5-7), and this loss of immune tolerance to insulin often leads to type 1 diabetes (8, 9). Immune tolerance to insulin is influenced by the human leukocyte antigen (HLA) DR4-DQ8 haplotype (8) and allelic variations in the *INSULIN* (*INS*) gene (10-12) via mechanisms involving thymic T cell deletion (13, 14).

Controlled exposure to antigen leads to protection against immune-mediated diseases such as childhood allergy (15) and animal models of autoimmunity (16). In type 1 diabetes, attempts have been made to reduce disease risk in individuals with established autoimmunity by administration of autoantigen orally (17, 18), intranasally (19, 20), intravenously, or subcutaneously (21, 22). Treatment-associated immune effects such as increases in antibody titers (20-22) and changes in CD4^+^ T cell responses to administered autoantigen were observed in some of these studies (20), indicating that administration could lead to immune modulation. Although none of these trials achieved their primary outcomes of diabetes prevention, beneficial treatment effects were observed in exploratory analyses of subgroups within the oral insulin immunotherapy trials (17, 18).

We reasoned that, similar to peanut allergy (15), the efficacy of antigen-specific immune therapy to prevent autoimmune disease would improve if treatment was started early in life and as a primary prevention therapy before individuals become autoantibody positive (23). There are several challenges in commencing autoantigen-specific immune-therapy in healthy autoantibody-negative children. Establishing safety is of utmost importance. Also important is the ability to identify the immune and other effects of exposure. We previously demonstrated that daily oral administration of high doses (67.5 mg) of insulin to children with a genetic risk of type 1 diabetes did not induce unwanted hypoglycemia and was associated with the induction of low affinity antibodies against insulin and insulin-responsive CD4^+^ T cell with features of regulation (24). These treatment-associated immune responses were not typical of autoimmune diabetes (8, 25). We, therefore, inferred that the treatment was likely to be safe and capable of inducing an immune response that might protect against the development of type 1 diabetes. While these earlier findings are an important proof of concept, they were obtained in a small number of children aged 2–7 years, which is after the period of greatest susceptibility to insulin autoimmunity and, therefore, late for primary prevention of islet autoimmunity. It was, therefore, necessary to assess safety and immune modulation in children aged less than 2 years. Here, we report the Pre-POInT-early randomized controlled trial in children aged 6 months–2 years, which represents the first intervention with autoantigen at this very early age and, therefore, uniquely analyses overall safety, immune responses and effects of exposure to exogenous autoantigen during peak susceptibility. Children in this age period undergo rapid growth, a transition from maternally derived immunity to acquired protection through exposure to vaccinations and infectious agents (26), and large changes in the immune repertoire and the gut microbiome. Daily exposure of the mucosal immune system to a key autoantigen in genetically susceptible children during this period presents a rare opportunity to assess the interplay between these factors in eliciting immune responses.

The Pre-POInT-early trial had four objectives. First, to determine the safety of daily oral insulin administration in very young children with high genetic susceptibility for type 1 diabetes; second, to determine whether the previously observed antibody and CD4^+^ T cell responses to oral insulin could be observed in younger children; third, to explore interactions between oral insulin therapy and *INS* genotype and microbiome; and fourth, to investigate immune changes and events that may influence autoimmunity during this period of high susceptibility.

## Results

### Participants

In total, 167 infants aged 6 months to 2 years with a first-degree family history of type 1 diabetes were screened (Figure 1A). Of these, 49 were eligible based on their HLA DRB1-DQA1-DQB1 genotype and the lack of antibodies to insulin (IAA), glutamic acid decarboxylase (GADA), insulinoma-associated antigen 2 (IA-2A), or zinc transporter-8 (ZnT8A). A susceptible *INS* genotype was not included in the eligibility criteria, but used to stratify the outcome analysis. Consent to participate was provided for 44 children (17 girls), who were randomized at a median age of 1.1 years (interquartile range [IQR], 0.8–1.7 years). There were no differences in baseline characteristics between the two groups (Table 1). The median exposure to oral insulin in 22 children was 12.0 months (cumulative exposure, 260.1 months; Supplemental Table 1) and to placebo in 22 children was 12.0 months (cumulative exposure, 252.5 months). All of the randomized children received at least one dose of their allocated treatment. The insulin dose was escalated from 7.5 to 22.5 mg and to 67.5 mg (Figure 1B), with cumulative exposure of 66.7, 67.2, and 126.2 months, respectively (Supplemental Table 1). The parents of one child in the placebo group withdrew their child from the study prior to attending the 3-month follow-up visit; no adverse event reporting or immune data were available from this child. Median adherence to the allocated treatments was 97.9% for placebo and 96.9% for oral insulin (Supplemental Table 1). Of 220 planned study visits, 212 (96.5%) were completed.

**Figure 1.**
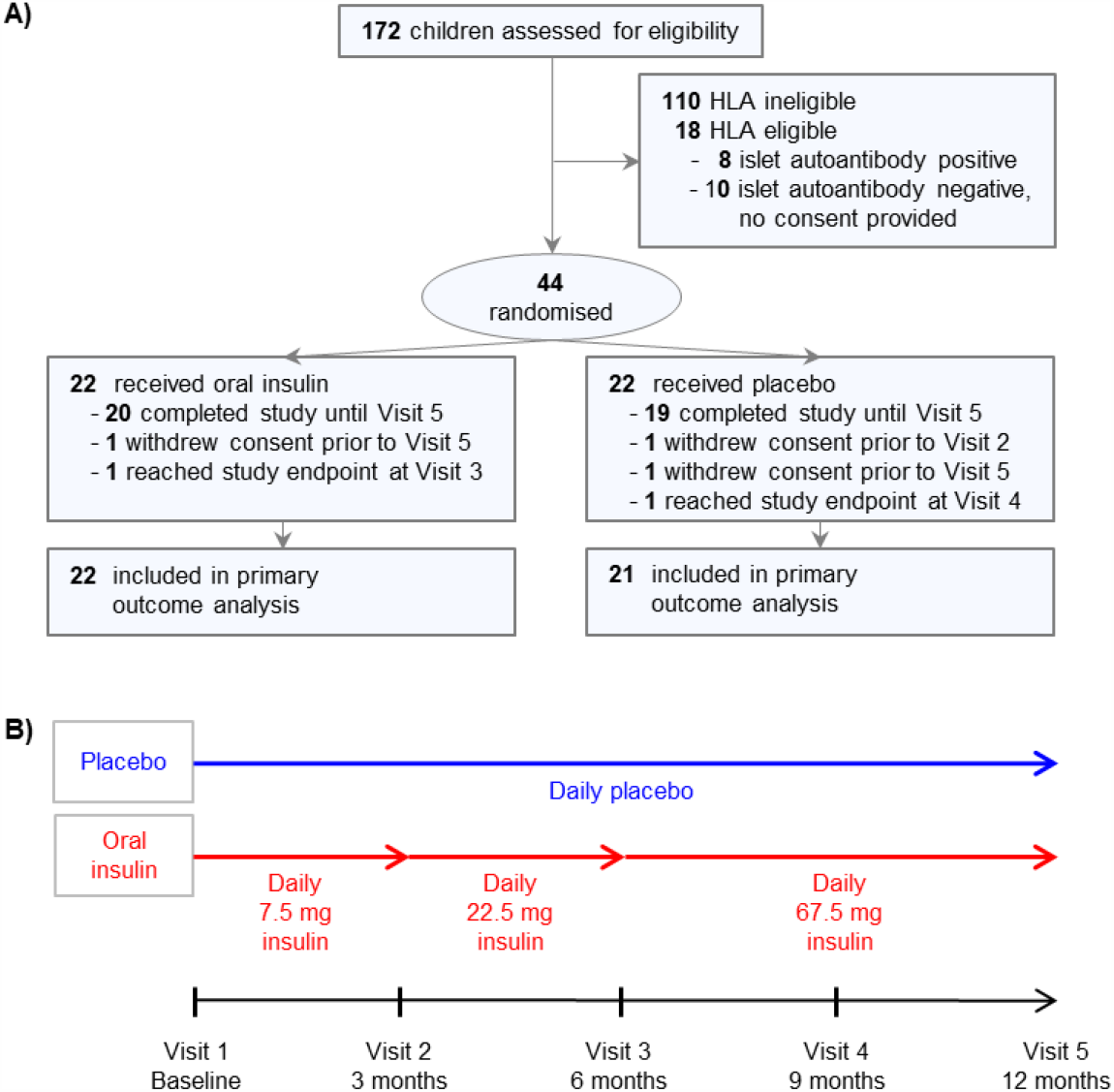
Schematics of participant disposition, design, and treatment groups. Disposition of the participants. All children had a first-degree family history of type 1 diabetes. Study end point was the development of persistent antibodies to GAD, IA-2 or ZnT8 (**A**). Study design and treatment groups (**B**).

**Table 1.**
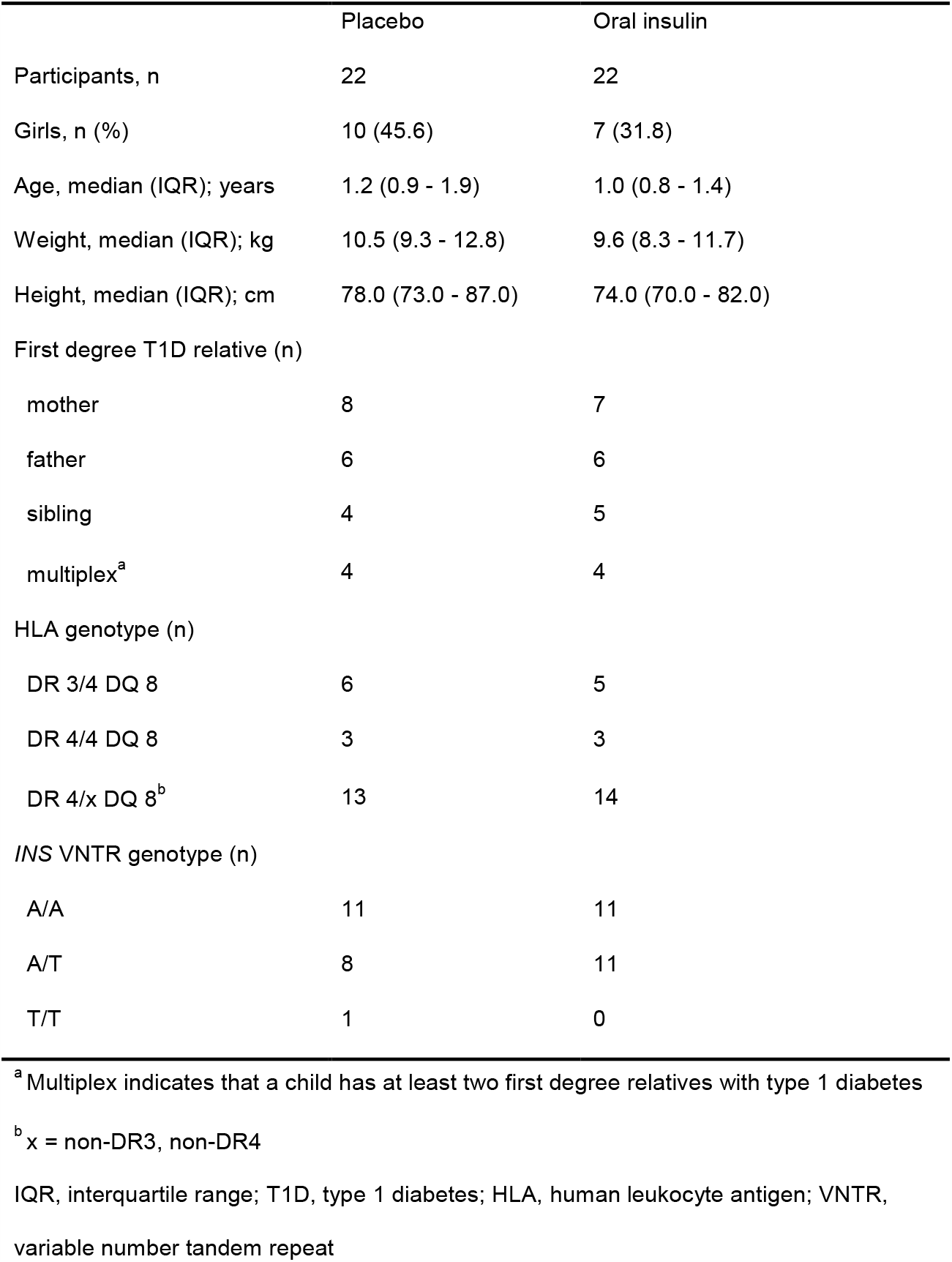
Baseline characteristics of children enrolled in the Pre-POInT-early study

### Glucose metabolism and laboratory findings

Oral insulin therapy was well tolerated with no evidence of treatment-related hypoglycemia. All blood glucose concentrations measured within 2 h after the first dose of placebo or oral insulin, or the first dose after each dose escalation were >50 mg/dL, except for one instance in a child in the placebo group (Figure 2A). There were no differences between the placebo and insulin group in blood glucose, insulin, or C-peptide values (Supplemental Table 2), the insulin/C-peptide ratio (Figure 2B), or the areas under the concentration–time curves for glucose, insulin, or C-peptide (Supplemental Table 2). Persistent islet autoantibodies against GAD, IA-2 or ZnT8 (study endpoint) developed 6.9 months after randomization in one child in the oral insulin group and 9.5 months in one child in the placebo group. There were no differences in blood counts and blood chemistry between the two groups (Supplemental Table 3).

**Figure 2.**
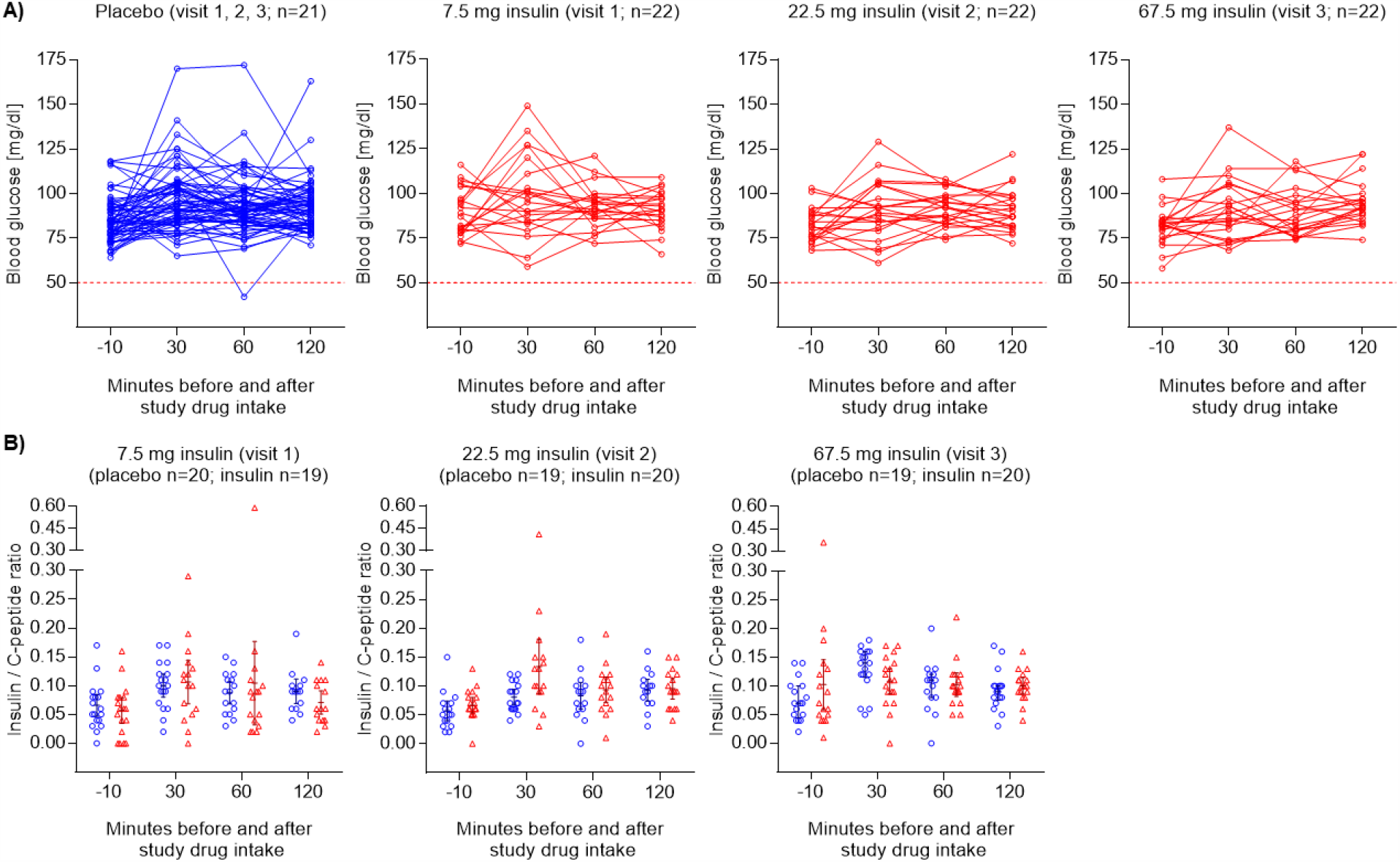
Blood glucose concentration and insulin/C-peptide ratio over time. Blood glucose concentrations (**A**) and insulin/C-peptide ratio (**B**) measured before and after study drug intake at baseline, 3 months, and 6 months. The dashed line indicates the threshold for hypoglycemia (50 mg/dL). The conversion factor for glucose values to millimoles per liter is mg/dL × 0.0555.

### Adverse events

A total of 114 adverse events were reported over a cumulative exposure period of 21.1 years in 21/21 children in the placebo group (5.64 events per year), and 181 adverse events were reported over a cumulative exposure period of 21.7 years in 22/22 children in the oral insulin group (8.38 events per year). The time to the first observed adverse event was not significantly different between the two groups (*P* = 0.39; log rank test). Infections were the most frequently reported adverse events (Supplemental Table 4). There was no difference in the severity of adverse events between the two groups. There were six serious adverse events, four in the oral insulin group and two in the placebo group, none of which were considered related to the study drug. By system organ class, the frequency of skin and subcutaneous tissue disorders was greater in the oral insulin group (12 events in 8 children) than in the placebo group (1 event in 1 child; *P* = 0.01; Supplemental Figure 1). These included diaper rash, erythema, eczema, pruritus, and urticaria (Supplemental Table 4). The overall frequency of skin and subcutaneous tissue disorders among all reported adverse events was 4.3% and all of these adverse events were classified as mild (grade 1) and resolved.

### Immunological response to daily oral insulin administration

The primary outcome was a positive antibody <u>or</u> CD4^+^ T cell response to insulin and was based on findings from the previous Pre-POInT trial (24). An antibody response was defined as an increase in serum IgG antibodies to insulin, salivary IgA antibodies to insulin, or serum IAA and a CD4^+^ T cell response was defined as a stimulation index (SI) above 3 that was more than 2-fold increased over baseline (see Methods). The primary outcome was observed in 26/43 (60.5%) children at 3 (*n* = 13), 6 (*n* = 9), or 9 months (*n* = 4) after randomization (Table 2), including 14/21 (66.7%; 95% confidence interval [CI] 45.4%–82.8%) in the placebo group and 12/22 (54.6%; 95% CI 34.7%–73.1%) in the oral insulin group (*P* = 0.54). Age-and sex-adjusted responses did not differ between the two groups (HR, 0.78; 95% CI, 0.36-1.70; *P* = 0.53).

**Table 2.** Primary outcome response to insulin

| Treatment group | Positive antibody<br>outcome<sup>a</sup> (n) | Positive CD4<sup>+</sup> T cell<br>outcome<sup>b</sup> (n) | Positive trial<br>outcome<sup>c</sup> (n) |
| --- | --- | --- | --- |
| All children (n=43) |  |  |  |
| Placebo (n=21) | 7 | 8 | 14 |
| Oral insulin (n=22) | 11 | 4 | 12 |
| *INS* AA genotype<br>(n=22) |  |  |  |
| Placebo (n=11) | 2 | 5 | 7 |
| Oral insulin (n=11) | 8 | 4 | 9 |

<sup>a</sup> A positive antibody outcome was defined as an increase in serum IgG antibodies to insulin, salivary IgA antibodies to insulin, or two-fold increase in serum IAA.
<sup>b</sup> A positive CD4<sup>+</sup> T cell outcome was defined as a CD4<sup>+</sup> T cell response to insulin (SI >3) that was more than 2-fold increased over baseline.
<sup>c</sup> A positive trial outcome was defined as a positive antibody outcome or positive CD4<sup>+</sup> T cell outcome.

The cumulative frequencies of antibody responses to insulin were 33.4% (95% CI 13.4%–53.2%) in the placebo group and 50.4% (95% CI 29.4%–71.4%) in the oral insulin group (*P* = 0.18; Figure 3A). Responses included an increased IgG binding to insulin (7 in placebo group and 9 in oral insulin group) and a salivary IgA response to insulin (1 in placebo group and 2 in oral insulin group). The cumulative frequencies of the positive CD4^+^ T cell responses to insulin were 38.5% (95% CI 18.5%–58.5%) in the placebo group and 18.4% (95% CI 2.4%–34.4%) in the oral insulin group (*P* = 0.15; Figure 3B). CD4^+^ T cell responses to insulin at 12 months were lower in the oral insulin group (median SI, 0.97; IQR, 0.71–1.21) than in the placebo group (median SI, 1.41; IQR, 0.94–2.2; *P* = 0.014; Figure 3C).

**Figure 3.**
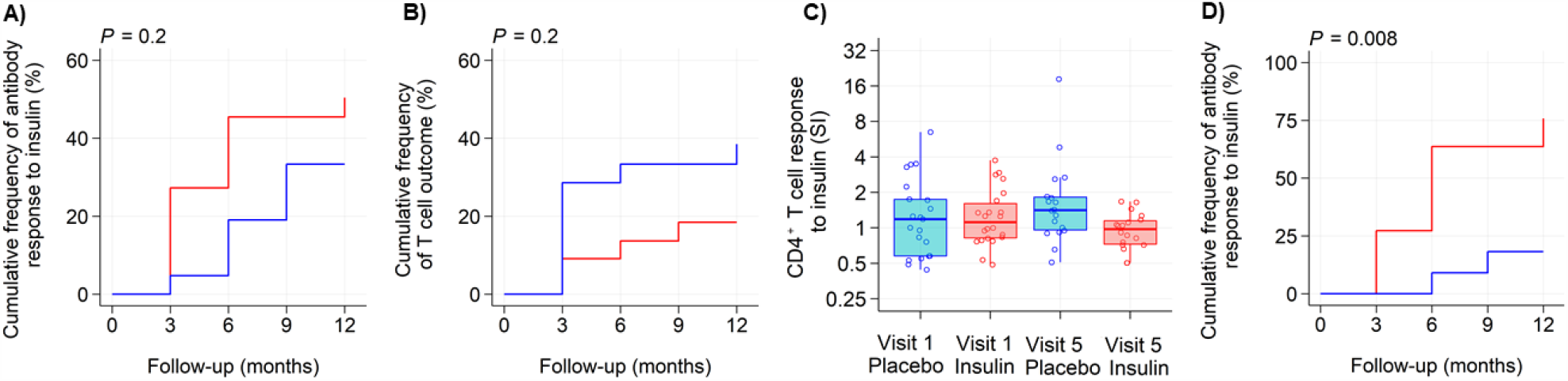
Responses to treatment and analysis of responses. Immune response to oral insulin or placebo. Kaplan–Meier analysis of a positive antibody response to insulin (**A**) and CD4^+^ T cell response to insulin (**B**) as defined by the primary outcome criteria in children who received placebo (blue line; n = 21) or oral insulin (red line; n = 22). The follow-up time is calculated from the first day of treatment (**A, B**). CD4^+^ T cell response to insulin calculated as the stimulation index relative to medium control at baseline (visit 1) and at 12 months (visit 5) in children who received placebo (blue circles; n = 21 at baseline, n = 18 at 12 months) or oral insulin (red circles; n = 22 at baseline, n = 18 at 12 months) (**C**). Kaplan–Meier analysis of a positive antibody response to insulin as defined by the primary outcome criteria in children with the *INS* AA genotype who received placebo (blue line; n= 11) or oral insulin (red line; n = 11) (**D**).

In conclusion, CD4^+^ T cell responses to insulin decreased in the oral insulin group, but no other differences in the primary outcome analysis of immune responses to insulin were observed between the placebo and oral insulin groups. As compared to the previous Pre-POInT study of children aged 2–7 years, the responses to insulin were more frequently observed in the placebo group of this study (66.7% vs 20%; *P* = 0.02) (24); the responses in the oral insulin groups, however, were similar in both studies (54.6% vs 60%). This suggests frequent activation of immune responses to insulin in very young genetically susceptible children in the study.

### The *INS* genotype was associated with immunological responses to treatment

Autoimunity against insulin is more frequent in children with the susceptible *INS* AA genotype (12, 27). However, there are no analyses of genetic modulation of responses to antigen-specific therapy. Therefore, the statistical analysis plan included comparisons of the frequency of immune responses to insulin between treatment groups in children with the susceptible *INS* AA genotype. An antibody response to insulin was observed in 10/22 children with the susceptible *INS* AA genotype, including 2/11 children in the placebo group (cumulative frequency at 12 months, 18.2%; 95% CI 0.1%–40.4%) and 8/11 children in the oral insulin group (75.8%; 95% CI 48.8%–99.9%; *P* = 0.0085; Figure 3D; Supplemental Table 5). The increased response in the oral insulin group remained significant after correction for multiple comparisons (*P* = 0.032; n = 4 comparisons). The majority of children with the susceptible *INS* AA genotype in the oral insulin group showed an antibody response by 6 months of treatment (cumulative frequency at 6 months, 63.6%, 95% CI 20.5–83.4%). An interaction between *INS* genotype and treatment was statistically tested using the Cox proportional hazards model and was observed for an antibody response (*P* = 0.024). Age was inversely associated with the antibody response in this model (*P* = 0.032). Age in the children with the susceptible *INS* AA genotype was not different between the placebo (median, 1.0 year) and oral insulin (median, 0.8 years) groups. Unlike the antibody responses, the frequency of T cell responses to insulin in children with the *INS* AA genotype was not different between the placebo group (5/11) and the oral insulin group (4/11; *P* > 0.99; Supplemental Table 5). These results indicate that oral insulin was effective in inducing an antibody response in very young children with a susceptible *INS* genotype in this study and, for the first time, show an immune modulating effect of the *INS* gene on oral insulin therapy analogous to that observed in the natural course of type 1 diabetes.

### Age, *INS* genotype, and treatment are associated with microbiome alterations

It is assumed that the potentially beneficial effects of oral exposure to antigen is via tolerogenic presentation in the oral mucosa. It is also possible that antigen may have beneficial effects on the mucosa. The administration of high doses of oral insulin to young children presented the possibility to assess effects on a changing microbiome and therefore, indirectly asses this. Of 116 stool samples collected from 42 participants, 115 were sequenced including 40 collected at baseline, 40 at 6 months, and 35 at 12 months follow-up. The average mapped reads per sample was 16,817 with a minimum (rarefication level) of 10,142. There was a marked relationship between the age of the children and the alpha and beta diversities of the microbiome. Bacterial richness (observed operational taxonomic units [OTUs]) and bacterial evenness (Shannon diversity) increased with age, and bacterial community metrics (unweighted Jaccard distance and weighted Bray–Curtis distance) converged with age (Figure 4, A-D). Similar findings were observed by whole genome sequencing (Supplemental Figure 2, A-D). There were significant changes in the abundances of several phyla and genera over time (Supplemental Figure 3, A and B).

**Figure 4.**
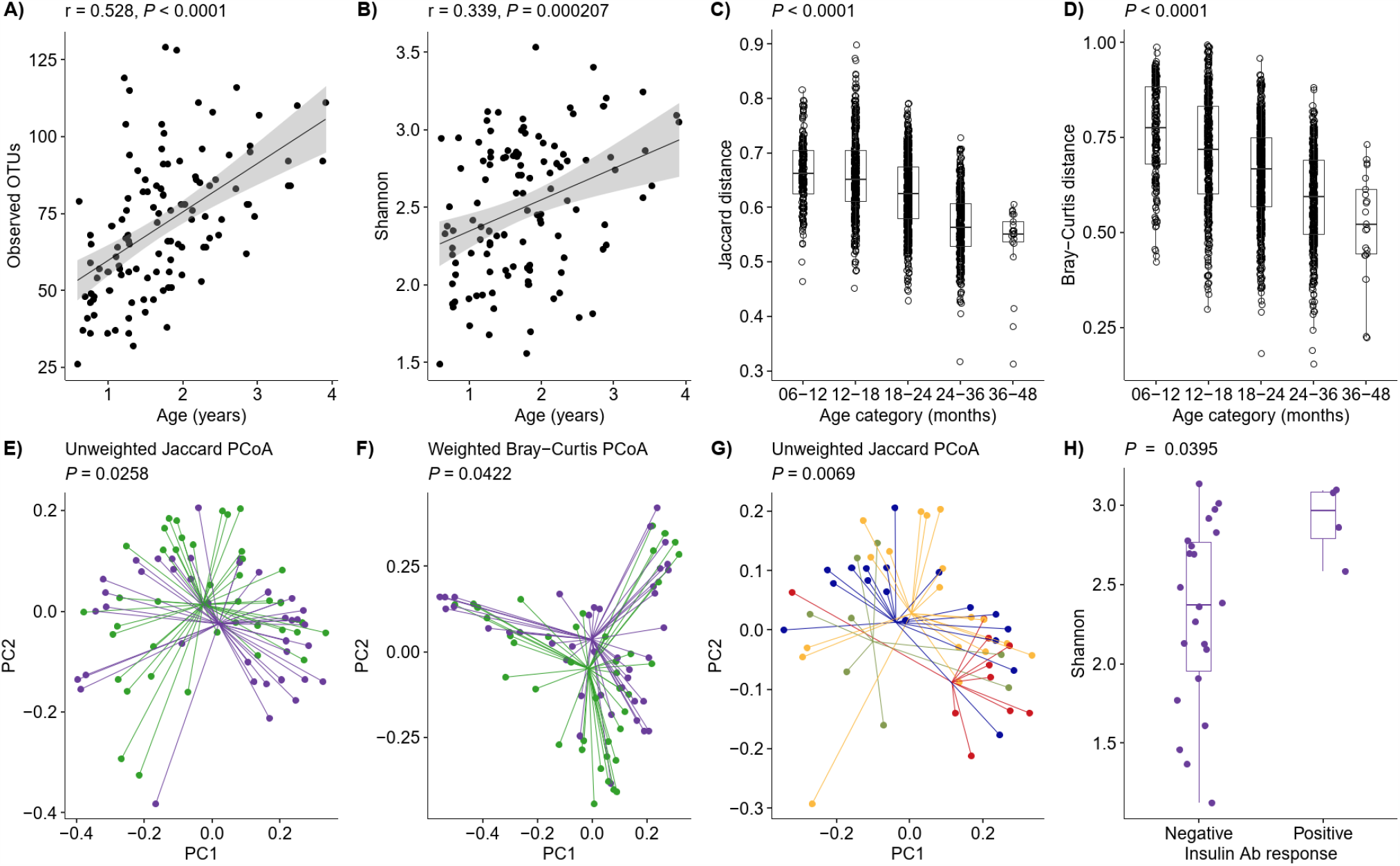
Microbiome alterations in relation to age, *INS* genotype, and treatment. Alpha diversity in relation to age over the time of study participation in samples from baseline (n = 40), 6 months (n = 40), and 12 months (n = 35); shown is richness (Observed OTU) (**A**) and evenness (Shannon) (**B**). Beta diversity in relation to age over time of study participation (baseline, 6 months, 12 months); shown is Jaccard distance (**C**) and Bray-Curtis distance (**D**). Each dot represents the distance between two samples within the age range (**C, D**). Beta diversity differences by PCoA (baseline, 6 months, 12 months); shown is Jaccard distance in children with the *INS* AA genotype (purple dots and lines) or the *INS* AT or TT genotype (green dots and lines) (**E**), Bray-Curtis distance in children with the *INS* AA genotype (purple circles and lines) or the *INS* AT or TT genotype (green circles and lines) (**F**), and Jaccard distance in children with the *INS* AA genotype who received placebo (blue dots and lines) or oral insulin (red dots and lines), and in children with the *INS* AT or TT genotype who received placebo (green dots and lines) or oral insulin (orange dots and lines) (**G**). Alpha diversity (Shannon) in children with the *INS* AA genotype who had a negative (n = 22 samples) or positive (n = 4 samples) antibody response to insulin (6 months, 12 months) (**H**). Unless indicated, the plots include both placebo and oral insulin-treated children. For treatment-related analyses, only post-baseline samples at 6 and 12 months were included.

The alpha and beta diversities of the microbiome did not differ between the oral insulin and placebo groups at 6- and 12-months follow-up. However, there were significant bacterial community differences between children with the *INS* AA genotype and children with the AT or TT genotypes, and between the treatment groups after stratification by *INS* genotype. The unweighted and weighted bacterial community metrics differed between children with the *INS* AA genotype and children with the AT or TT genotypes (unweighted Jaccard distance *P* = 0.025; weighted Bray–Curtis distance *P* = 0.042; Figure 4, E and F). The relative abundance of *Bacteroides dorei* was increased in children with the *INS* AA genotype (6.2%) versus children with the AT or TT genotypes (0.4%; *P* = 0.002; Supplemental Figure 4A). The unweighted Jaccard distance in children with the *INS* AA genotype who received oral insulin differed as compared to the placebo group and to children with the AT or TT genotypes (*P* = 0.0069; Figure 4G). Subtle differences in alpha diversity included a more pronounced age-related increase in bacterial richness, as measured by the observed OTUs, in children with the *INS* AA genotype in the oral insulin group as compared with the placebo group (Supplemental Figure 4, B and C). There was also a potential increase in bacterial evenness (Shannon diversity) observed among children with the *INS* AA genotype who showed a positive antibody response to insulin as compared with children who showed no antibody response (*P* = 0.04; Figure 4H). The findings identify an association between early microbiome development and the *INS* gene and suggest that *INS* gene effects on immune responses to insulin may in part be acting via the gut.

### Type 1 interferon profiles were frequent and associated with inflammatory markers and events

In addition to examining treatment-related effects, the trial provided the opportunity to assess multiple immune parameters in children during a critical age of exposure to vaccination, infection, and susceptibility to insulin autoimmunity. We found cell populations that were constant over the 12-month follow-up within individuals but showed strong inter-individual differences (Treg, CD8^+^ T cells; Supplemental Figure 5, A and B); other cell populations varied across measurements within children (activated CD8^+^ T cells, intermediate monocytes; Supplemental Figure 5, C and D). Age was strongly correlated with the frequency of peripheral blood mononuclear cell populations (Supplemental Figure 6, A and B).

Type 1 interferon signatures have been linked to type 1 diabetes (28). We frequently observed samples with CD169 (Siglec-1) expression on monocytes (59/208, 28.5%; Figure 5, A and B) in the children, an indication of an ongoing type 1 interferon response (28). This feature was observed in 32/43 (74%) children and was found on multiple samples in 19 (44%) children. CD169 expression on monocytes was associated with the relative frequency of the inflammatory type intermediate (CD14^++^CD16^+^) monocytes (*r* = 0.52; *P* <0.0001; Figure 5C), and both of these features were associated with an increased frequency of activated (CD69^+^) T cells (CD169^+^ monocytes, r = 0.34, *P* <0.0001; intermediate monocytes, r = 0.65, *P* <0.0001). Monocyte CD169 expression was also correlated with the levels of the proinflammatory plasma proteins CXCL10 (r = 0.49; *P* = 0.0002), IL-6 (r = 0.42; *P* = 0.0038), and IFNγ (r = 0.39; *P* = 0.0095; Supplemental Table 6), and associated with decreased gut microbiome richness (*P* = 0.033; Supplemental Figure 7, A and B) and younger age (*P* = 0.014; Supplemental Figure 7C). To determine whether the monocyte CD169 expression corresponded to potential inflammatory episodes in the children, we examined the trial adverse events in the two week period prior to sample collection and observed more adverse events when samples were monocyte CD169^+^ (20/49, 40.8%) than when samples were CD169^−^ (21/118, 17.8%; *P* = 0.0028). Therefore, CD169 expression on monocytes marked inflammatory episodes in children.

**Figure 5.**
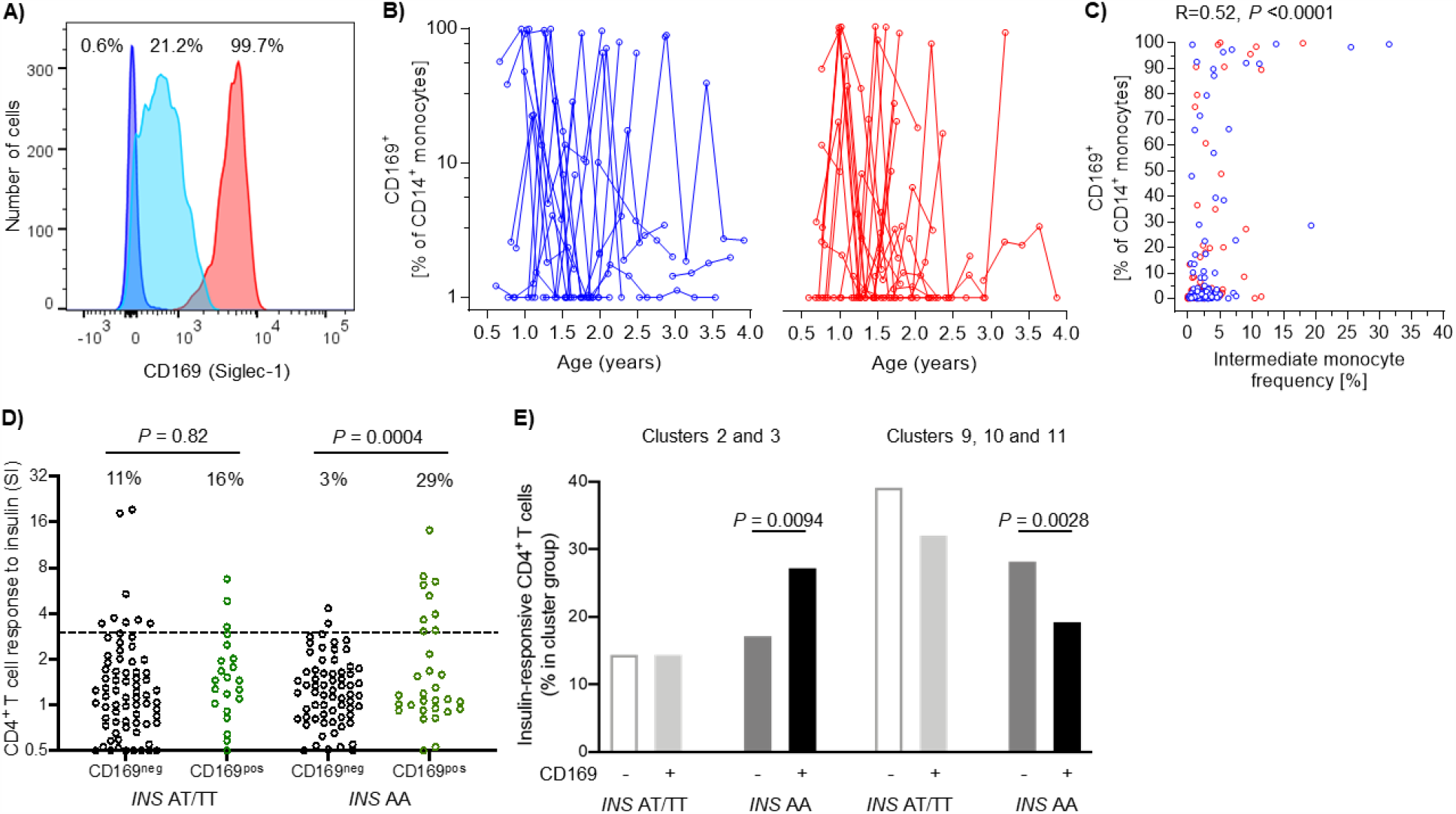
T cell responses to insulin in relation to *INS* genotype and monocyte CD169 expression. Monocyte CD169 expression and CD4^+^ T cell responses to insulin over the time of study participation (baseline, 3, 6, 9, 12 months) in all study participants (**A-D**). Representative flow cytometry histograms of CD169 staining intensity on monocytes. Shown are three samples with low (blue; 0.6% positive cells), moderate (light blue, 21.2% positive cells) and high monocyte CD169 expression (red, 99.7% positive cells). A threshold of >5% positive monocytes was used as the threshold for defining monocyte CD169^+^ samples (**A**). Percentage of CD169^+^ cells out of CD14^+^ monocytes in relation to age over the time of study participation in children who received placebo (blue lines; n = 21) or oral insulin (red lines; n = 22) (**B**). Correlation between the frequency of CD169^+^ monocytes and the frequency of intermediate monocytes in peripheral blood from children who received placebo (blue circles; n = 102 samples) or oral insulin (red circles; n = 104 samples) (**C**). CD4^+^ T cell responses to insulin (SI) in samples from 20 children with the *INS* AT or TT genotype and 22 children with the *INS* AA genotype and stratified by monocyte CD169 expression as negative (CD169^neg^, black circles; n = 148 samples) or positive (CD169^pos^, green circles; n = 58 samples) in all study visits (**D**). The frequencies of insulin-responsive CD4^+^ T cells (n = 1036 cells from 22 samples) in the Th1/Th21-like cell clusters 2 and 3 (left) and the Treg-like clusters 9, 10, 11 (right) according to whether cells were from children with the *INS* AT/TT (n = 550 cells) or *INS* AA (n = 486 cells) genotype and samples that were monocyte CD169 negative (white and dark gray bars, n = 559 cells) or positive (light gray and black bars, n = 477 cells) (**E**).

### Type 1 interferon profiles interacted with *INS* genotype to promote CD4^+^ T cell responses to insulin

Overall, an inflammatory response with type 1 interferon features was surprisingly frequent in these young genetically susceptible children. We, therefore, speculated that this may play a role in the pathogenesis of insulin autoimmunity. Strikingly, we found an association between CD169 expression and positive CD4^+^ T cell responses to insulin (SI > 3; 12/50 CD169^+^ vs 10/146 CD169^−^; *P* = 0.0026), suggesting that the *in vitro* assay may be affected by the inflammatory state of the child. The *INS* gene modified this association, which was only observed in children with the susceptible genotype (Figure 5D). Among children with the *INS* AA genotype, CD4^+^ T cell responses to insulin were observed in 9/31 (29%) CD169^+^ samples versus 2/68 (3%) CD169^−^ samples (*P* = 0.0004). A similar relationship between monocyte CD169 expression and T cell response to insulin in children with the *INS* AA genotype was observed for CD8^+^ T cells (Supplemental Figure 8). Monocyte CD169 expression was not associated with antibody responses to insulin.

To determine whether the phenotypes of the *in vitro* responses were also influenced by the inflammatory state of the child, we examined single cell transcription profiles of the insulin-responsive CD4^+^ T cells in relation to monocyte CD169 expression. The expression of 76 genes was analyzed in 1036 insulin-responsive CD4^+^ T cells from samples with a >3 SI response to insulin. The profiles were distributed in 11 cell clusters (Supplemental Figure 9A), including two clusters with features of Th1/Th21-like T cells (*IFNG, IL-21*; clusters 2 and 3; n = 184 cells), and three clusters with features of Tregs (*FOXP3*, low *CD127* and low cytokine expression; clusters 9, 10, and 11; n = 313 cells; Supplemental Figure 9B). The distribution across clusters differed between cells from CD169^+^ and CD169^−^ samples (*P* = 1.5 × 10^− 10^), between cells from children with the *INS* AA genotype and the AT or TT genotypes (*P* = 4.3 × 10^− 7^), and between the CD169^+^ and CD169^−^ samples from children with the *INS* AA genotype (*P* = 0.0003), but not among samples from children with the AT or TT genotypes (*P* = 0.060). Among children with the *INS* AA genotype, the CD169^+^ samples contained a higher proportion of insulin-responsive cells in the Th1/Th21-like clusters (88/323; 27.2%) than the CD169^−^ samples (28/163, 17.2%; *P* = 0.0094) and a lower proportion of insulin-responsive cells in the Treg-like clusters (67/323, 19.4% vs. 46/163, 28.2%; *P* = 0.0028; Figure 5E). Altogether, these findings suggest that *in vitro* presentation of insulin to T cells by antigen presenting cells will more likely result in a productive Th1/Th21-like T cell response if the cells are from a child with a susceptible *INS* genotype and in an active inflammatory state.

## Discussion

The Pre-POInT-early study is the first to expose very young genetically at risk children to exogenous autoantigen at an age of peak susceptibility to autoimmunity. It demonstrated that daily oral administration of up to 67.5 mg insulin to healthy, genetically at-risk, islet autoantibody-negative children at 6 months to 2 years of age was well tolerated without signs of hypoglycemia. The study failed to meet its primary outcome of immune efficacy defined by the findings in older children (24) as a composite of antibody and T cell responses to insulin. Treatment effects were, however, found for CD4^+^ T cell responses to insulin and, in particular, in sub-group analyses of children with the susceptible *INS* genotype, which included antibody responses to insulin and differences in the microbiome between children in the oral insulin and placebo groups. The *INS* gene-associated effects provide strong evidence that this gene generally affects immune response to exogenous and endogenous insulin and highlight how disease susceptibility genes may have unexpected mechanisms of action such as modifying microbiome. The study also revealed remarkably frequent treatment-independent inflammatory episodes with features of type 1 interferon responses in the participants. These inflammatory episodes influenced insulin-directed T cell responses, again in an *INS* gene associated manner, providing a potential mechanism for the high incidence of islet autoimmunity in early childhood.

Although hypoglycemia was not previously reported during treatment with oral insulin (17, 18, 24), children <2 years of age have not been exposed to oral insulin. Therefore, the absence of hypoglycemia at any of the tested doses with a cumulative exposure of >21 years is an important safety outcome. We also found no differences in glucose, insulin, and C-peptide over a 2-h period after administration of insulin compared with administration of a placebo. To our knowledge, this is the first study to include comprehensive metabolic measures upon administration of oral insulin in all participating children. These data indicate that oral insulin is unlikely to enter the blood stream, a conclusion that was important for initiating the POInT phase 2b trial in 4–6-month-old infants (29). Of note, the induction of tolerance by oral antigen is thought to be via antigen uptake in the oral and/or gut mucosa and does not require entry into the blood stream. As in the Diabetes Prevention Trial-Type 1 (DPT-1) (17), TrialNet (18), and Pre-POInT (24) trials, we observed no signs of allergy or intolerance to orally administered insulin. The frequency of adverse events was not increased in the oral insulin group, except for skin and subcutaneous tissue disorders. This was not observed in larger secondary prevention DPT-1 (17) and TrialNet (18) trials, where children from 3 years of age were treated with a daily dose of 7.5 mg oral insulin. It is possible that our finding was due to the exposure of children at ≤3 years of age or the use of higher oral insulin doses that may increase the likelihood of skin exposure to study drug. The overall frequency of skin and subcutaneous tissue disorders among all reported adverse events (4.3%) is comparable with that in TrialNet (7.7%) (18). Unlike some of the previous studies, our study included mandatory reporting of grade 1 (mild) events. All skin and subcutaneous tissue adverse events in our study were classified as mild, resolved during course of the study, and were not correlated with other blood chemistry measurements or inflammatory markers.

In addition to establishing safety, our objective was to find evidence for a treatment-induced immune response, which included IgG binding to insulin, salivary IgA against insulin, and CD4^+^ T cell responses to insulin. The study design and sample size were based on results from the previous Pre-POInT study, which enrolled children at 2–7 years of age with greater genetic risk (24). Using the same outcomes and measurement methods, we observed a higher overall reactivity to insulin in the placebo group in this study and the primary outcome was not met. The younger age of the children is a major difference of the current study and is likely to contribute to the higher observed frequency of immune responses to insulin in the placebo group. Evidence for this include the association between the antibody responses to insulin and younger age, and the correlation between age and T cell and monocyte subset compositions, with the latter being associated with the T cell responses to insulin.

Limitations of the study include the short follow-up period on relatively few children, which prevented us from assessing the efficacy of treatment in preventing islet autoimmunity or type 1 diabetes. A strength of the study is that, despite the challenge of obtaining blood samples from young children, adherence to the study protocol was high with comprehensive sample and data collection. This allowed us to collect extensive mechanistic data under the stringent conditions of a randomized controlled trial, in which all data with the exception of the plasma inflammatory markers were collected centrally before unblinding. These data included deep phenotyping of the immune responses and microbiome during early childhood and provided new insights into how oral insulin perturbs the immune system and disease mechanism. As there were no previous data to justify their inclusion in primary analyses, a number of these findings were based on exploratory and sub-group analyses, and, therefore, require validation in subsequent studies such as the POInT trial (29).

An important finding was that the *INS* genotype influenced some of the immune efficacy measures. In particular, we observed a robust effect of oral insulin on the antibody response in children with the susceptible *INS* AA genotype, suggesting that the *INS* gene modifies the likelihood of the immune system responding to oral insulin. The response was observed by 6 months of treatment, corresponding to a 22.5 mg dose, which is lower than in the previous study (24) and may reflect the lower body weight of children in the present study. Genetic susceptibility criteria in the previous study were more stringent and it is likely that the majority of participants had the susceptible AA genotype. We also discovered differences in the microbiome composition between children with susceptible and non-susceptible *INS* genotypes and after oral insulin treatment in children with the susceptible *INS* AA genotype. This included differences in bacterial diversity and richness, and an increased abundance of *Bacteroides dorei* in children with the susceptible *INS* genotype, a finding that is consistent with, and may explain, the increased abundance of *Bacteroides dorei* in children who developed type 1 diabetes in a Finnish study (30). Therefore, oral insulin was associated with an antibody response and changes in the gut microbiota and both of these effects were modified by *INS*. A question emerging from these findings is whether these were independent associations or whether the effects of oral insulin on the microbiome increased the likelihood of an antibody response. The increased alpha diversity in children treated with oral insulin who showed an antibody response compared with those without a response is consistent with a microbiome-mediated treatment effect.

Unlike the antibody outcome, the *in vitro* T cell responses to insulin were not associated with treatment after stratification by the *INS* genotype. The T cell responses were, however, strongly associated with monocyte CD169 expression, providing new insights into disease pathogenesis. Monocyte CD169 expression is a sensitive marker of a type 1 interferon signature, which increases before islet autoantibody seroconversion in young children and is associated with respiratory infection (28). CD169^+^ samples were surprisingly frequent and observed in the majority of children. They were also associated with recent adverse events, younger age, and several other inflammatory markers. The frequent inflammatory responses in this age period may reflect increased exposure to infection, vaccines, diet, or other inflammatory conditions, and/or a greater inflammatory response in young children that may be downstream of epigenetic and metabolic reprogramming (31-34). Early infection is associated with islet autoimmunity (35-37) and type 1 diabetes (38). Thus, our findings that *in vitro* T cell responses to insulin were more likely to occur in CD169^+^ samples in children with the susceptible *INS* AA genotype may be relevant to the mechanism of insulin autoimmunity. We believe that our results do not reflect the presence of *in vivo* primed T cells, but rather a heightened ability of the CD169^+^ monocytes to activate naive T cells in the *in vitro* assay. Children with the susceptible *INS* genotype are expected to have more peripheral insulin-autoreactive T cells (13, 14). Extending our *in vitro* findings, we suggest that a type 1 interferon response to infection in antigen-presenting cells *in vivo* further increases the likelihood of activating these T cells and eventually leads to insulin autoimmunity. The observation that the insulin-responsive cells from the CD169^+^ samples contained more Th1/Th21 cells and fewer Tregs supports this hypothesis, and may also explain our previous finding of proinflammatory, proinsulin-responsive T cells in infants who later developed islet autoimmunity (25). The requirement of danger signals for the activation of naive T cells is well documented (39) and findings in the NOD mice with respect to type 1 interferon receptor generally support a role of innate immune pathways, activated macrophages and microbiome in autoimmune diabetes (40-42). However, to our knowledge, this is the first demonstration of a relationship between the innate immune status and the T cell response to an antigen in unmanipulated samples obtained from children.

Overall, this study demonstrated safety for high dose oral insulin administration in young children. Immune response analyses provided evidence that orally administered insulin acted on the immune system of very young genetically susceptible children and that the immune response to oral insulin was modified by the *INS* gene as previously demonstrated for autoantigen. We, therefore, advocate that ongoing and future trials that include insulin or peptides of proinsulin as antigen specific immunotherapy (43, 44) should incorporate stratification by *INS* genotype into their study design and analyses. The efficacy of oral insulin to prevent islet autoimmunity is currently under investigation (29).

## Methods

### Participants

The Pre-POInT-early study was a randomized, placebo-controlled, double blind, single-center, pilot phase II clinical study (Clinicaltrials.gov NCT02547519). Participants were recruited between December 2015 and December 2016. Follow-up visits were completed in December 2017. Children were eligible if they were aged 6 months to 2.99 years, seronegative for IAA, GADA, IA-2A, and ZnT8A, and at high genetic risk of developing type 1 diabetes. High genetic risk was defined by a first-degree relative with type 1 diabetes diagnosed before 40 years old and a HLA genotype that included the HLA DR4-DQB1*0302 or HLA DR4-DQB1*0304 haplotypes, and did not include one of the following alleles or haplotypes: DR 11, DR 12, DQB1*0602, DR7-DQB1*0303, DR14-DQB1*0503.

### Randomization and masking

A computer-generated randomization list was prepared with an allocation ratio of 1:1 (placebo to oral insulin) using a web-based system (https://wwwapp.ibe.med.uni-muenchen.de/randoulette/). All investigators and participants were masked to the treatment allocation. Unblinding was not necessary during the study.

### Intervention

Insulin crystals were provided by Lilly Pharmaceuticals (Indianapolis, Indiana). The investigational products (insulin and placebo) were manufactured as identical capsules containing either insulin crystals (7.5, 22.5, or 67.5 mg) in microcrystalline cellulose (total capsule content 200 mg) or 200 mg microcrystalline cellulose placebo by InPhaSol, Apotheke des Universitätsklinikums Heidelberg, Germany. The drug packages were sequentially numbered according to the randomly allocated treatment. Parents were instructed to sprinkle the contents of one capsule onto one teaspoon of food (e.g. yogurt, breast milk, or commercial baby food) for administration once daily.

Children were randomized 1:1 to receive oral insulin or placebo for a period of 12 months. Children in the oral insulin group received 7.5.mg of insulin for 3 months, then 22.5 mg for 3 months, and finally 67.5 mg for 6 months (Figure 1B). Follow-up visits were scheduled at 3, 6, 9, and 12 months after starting treatment.

### Procedures

Blood samples and saliva were collected at each visit to determine islet autoantibodies, immune responses to insulin, lymphocyte and monocyte subsets, and salivary IgA antibodies to insulin. At the baseline, 3-month, and 6-month visits, blood samples were collected before (− 10 min) and 30, 60, 90, and 120 min after study drug intake to measure blood glucose, insulin, and C-peptide. Blood cell counts, liver function, electrolytes, IgE, and plasma markers of inflammation were measured at baseline (visit 1) and 12 months (visit 5). Stool samples were collected to assess the microbiome at baseline, 6 months, and 12 months. Medication adherence was assessed by family self-reporting of daily capsule administration using adherence sheets. Children reached study endpoint and stopped treatment if they developed persistent autoantibodies (GADA, IA-2A, or ZnT8A) or clinical diabetes. The development of IAA was not considered as an endpoint as it was considered a potential response to oral insulin. The final statistical analysis plan of the study was signed on July 3, 2018. All data except the plasma markers of inflammation were submitted before the data hardlock which was signed on July 18, 2018: Data was unblinded by the independent study statistician on July 24, 2018.

### Measurements related to safety

Serum insulin and C-peptide concentrations were measured by fluorescence enzyme immunoassays using an automated immunoassay analyzer (AIA-360, Tosoh Bioscience Inc., South San Francisco, CA). The families were instructed to note any symptoms of hypoglycemic events such as trembling, sweating or impaired consciousness after study drug intake. Hypoglycemia was defined as a blood glucose level <50 mg/dL (2.78 mmol/L). IgE against insulin was measured using a radiobinding assay (45). Islet autoantibodies (GADA, IA-2A, and ZnT8A) were measured using harmonized radiobinding methods (46,47). Throughout the study, the investigators recorded any adverse events using an adverse event clinical report form, regardless of the event’s severity or relation to the study drug or study procedure.

### Measurements of immunological response

The primary immune efficacy outcome was an immune response to insulin, defined as an increase in any one or more of the following: serum IgG antibodies to insulin, salivary IgA antibodies to insulin, serum IAA, or a CD4^+^ T cell response to insulin. Additional outcomes included the antibody and CD4^+^ T cell responses in children with the *INS* AA genotype, the gene expression profile CD4^+^ T cells responding to insulin, T lymphocyte and monocyte subset frequencies, stool microbiome, and plasma inflammatory markers.

Insulin autoantibody levels were measured using a competitive radiobinding assay (48, 49). A positive response was defined as a value of ≥1.5 and a ≥2-fold increase from baseline. IgG binding to insulin was measured by a non-competitive radiobinding assay with protein-G capture of IgG (50). A positive response was defined as an increase of >10 counts/min from the baseline value. Salivary IgA binding to insulin was measured using a radiobinding assay as previously described (24). A positive response was defined as an increase of ≥3-fold from baseline.

CD4^+^ T cell antigen responses were measured using stored frozen peripheral blood mononuclear cells (PBMCs). Responses were measured using a dye (Cell Proliferation Dye eFluor 670, eBioscience, San Diego, CAL) dilution assay, quantifying proliferation (eFluor670dim cells) and activation (CD25^+^) after 5 days of culture without or with the antigen insulin (50 μg/mL, Lilly Pharmaceuticals) as previously described (24) (Supplemental Figure 10). The assay included a median of 12 wells containing 200,000 eFlour670 dye-labelled cells in medium plus insulin and 6 wells with cells and medium alone. The SI was calculated as the number of CD4^+^ eFluor670^dim^CD25^+^ cells per 50,000 acquired live CD4^+^ T cells in all wells containing insulin relative to the number of CD4^+^ eFluor670^dim^CD25^+^ cells per 50,000 acquired live CD4^+^ T cells in all wells containing medium alone. A positive sample was defined as an SI of >3. A positive T cell outcome was defined as a positive sample and an increase in SI of >2-fold at any follow-up visit relative to the baseline value. CD8^+^ T cell proliferation responses to insulin were also measured in the same assay by gating on CD8^+^CD4^−^ T cells (Supplemental Figure 10).

### *INS* genotyping

Genomic DNA was amplified using primers (forward: 5’-GGTCTGTTCCAAGGGCCTTT-3’; biotinylated reverse: 5’-ATGGCAGAAGGACAGTGATCTGG-3’) targeting rs689 of *INS* and the SsoFast EvaGreen Supermix on a CFX96 system (Bio-Rad, Hercules, CA) according to manufacturer’s protocol. Subsequently, genotyping of rs689 was performed by pyrosequencing on a PyroMark Q48 Autoprep using a sequencing primer (5’-CTCAGCCCTGCCTGT-3’) and PyroMark Q48 Advanced Reagents (Qiagen, Hilden, Germany) according to manufacturer’s protocol. Primer design and SNP analysis was carried out using the PyroMark Assay Design 2.0 and PyroMark Q48 Autoprep 2.4.2 Software (Qiagen), respectively.

### Phenotyping of lymphocytes and monocytes

Freshly isolated PBMCs (2.5 × 10^5^ cells) were incubated for 1 min at room temperature with Fc receptor blocking reagent (Miltenyi Biotec, Bergisch Gladbach, Germany). Cell surface markers were stained for 20 min at 4 °C in phosphate-buffered saline (PBS) without Ca^2+^ and Mg^2+^ (PBS^− /−^; Gibco) containing 0.5% bovine serum albumin using the following mouse anti-human monoclonal antibodies: anti-CD3 Alexa Fluor 700 (clone HIT3α; BioLegend, San Diego, CAL), anti-CD4 Brilliant Violet 510 (SK3; BD Biosciences, San Jose, CAL), anti-CD8a Brilliant Violet 605 (RPA-T8), anti-CD14 Pacific Blue (HCD14), anti-CD16 PerCP-Cy5.5 (3G8; all BioLegend), anti-CD25 PE (M-A251; BD Biosciences), anti-CD45RA PE-Cy5 (HI100; BioLegend), anti-CD69 fluorescein isothiocyanate (FN50; BD Biosciences), anti-CD127 PE-Cy7 (A019D5) and anti-CD169 Alexa Fluor 647 (7-239; both BioLegend). Cells were washed twice in PBS^− /−^ and stained for 20 min at room temperature with Zombie NIR (BioLegend) to evaluate cell viability. PBMCs were fixed with 1.5% formalin in PBS^− /−^ and analyzed within 24 h on a flow cytometer (LSR Fortessa, Becton Dickinson, Franklin Lakes, NJ) using FACSDiva acquisition software (Version 7.0; BD Biosciences). FlowJo software (Version 10; TreeStar Inc., Ashland, OR) was used to analyze lymphocyte and monocyte subsets.

### Single-cell gene expression profiling

CD4^+^ T cells that had proliferated, as determined by eFluor® 670 dilution, and displayed CD25 upregulation were identified as responding cells and were single-cell-sorted directly into 96-well microplates containing 5 μL of PBS prepared with diethylpyrocarbonate-treated water. For samples that had an SI against insulin above 3, cells were processed for gene expression. cDNA was synthesized directly from cells using qScript(tm) cDNA Supermix (Quanta Biosciences, Gaithersburg, MD). Total cDNA was pre-amplified for 18 cycles, with 1 cycle of denaturation at 95 °C for 1 min, followed by cycling at 95 °C for 15 s, 60 °C for 1 min, and 72 °C for 1.5 min, followed by one cycle of 72 °C for 7 min, with TATAA GrandMaster Mix (TATAA Biocenter, Göteborg, Sweden) in the presence of 76 primer pairs at a final volume of 35 μL. Then, 10 μL of preamplified DNA was treated with 1.2 units of exonuclease I. To quantify gene expression, real-time PCR was performed on the BioMark(tm) HD System (Fluidigm Corporation, South San Francisco, CA) using the 96.96 Dynamic Array IFC according to the GE 96 × 96 Fast PCR+ Melt protocol with SsoFast EvaGreen Supermix containing Low ROX (Bio-Rad) and 5 μM of primers in each assay. The primers and target genes are listed in Table S7. Raw data were analyzed using Fluidigm Real-Time PCR analysis software and GenEx Pro 5.3.6 Software (MultiD, Göteborg, Sweden). Additional data analysis was done using KNIME 2.5.2 software. Analysis of multivariate gene expression patterns was performed by Uniform Manifold Approximation and Projection for Dimension Reduction (UMAP) (51) and unsupervised WARD hierarchical clustering (hclust) on the pre-processed Ct values. For pre-processing, a linear model was used to correct for potential confounding effects, which can mask relevant biological variability (52). In brief, batch effects (dummy coding for each plate/batch) were modeled jointly with dose effects by regressing out the effect of plates on each individual gene while controlling for dose in order to obtain a corrected gene expression dataset.

### Stool microbiome

Stool samples were collected at home at the day of the visit or within two days before or after the visit at baseline, 6 months, and 12 months, into tubes containing ethanol that were provided to the parents. The samples were brought to the visit, aliquoted, and stored at − 80 °C. Alternatively, samples could be sent to the central laboratory with guaranteed delivery within 24 h. The bacterial component of the microbiome in each stool sample was analyzed by 16*S* rRNA gene compositional analysis, as previously described (26). To generate 16*S* rDNA data, genomic bacterial DNA was extracted from the samples using MO BIO PowerMag Soil DNA Isolation Kit (Qiagen). The 16*S* rDNA V4 region was amplified by PCR and sequenced in the MiSeq platform (Illumina) using the 2 × 250 bp paired-end protocol. The primers used for amplification contained adapters for MiSeq sequencing and dual-index barcodes so that the PCR products may be pooled and sequenced directly (53), targeting at least 10,000 reads per sample. The standard pipeline for processing and analyzing the 16S rDNA gene data incorporated phylogenetic and alignment-based approaches to maximize data resolution. The read pairs were demultiplexed based on the unique molecular barcodes, and reads were merged using USEARCH v7.0.1001 (54). 16S rRNA gene sequences were assigned into Operational Taxonomic Units (OTUs) or phylotypes at a similarity cutoff value of 97% using the UPARSE pipeline. Abundances were recovered by mapping the demultiplexed reads to the UPARSE OTUs.

A subset of samples was selected for metagenomic whole genome shotgun (WGS) sequencing for deeper characterization. Metagenomic WGS sequencing utilized the same extracted bacterial genomic DNA used for 16S rDNA compositional analysis. For WGS, individual libraries constructed from each sample were loaded into the HiSeq platform (Illumina) and sequenced using the 2×100 bp pair-end read protocol. The process of quality filtering, trimming and demultiplexing was carried out by in-house pipeline developed by assembling a number of publicly available tools such as Casava v1.8.3 (Illumina) for the generation of fastqs, Trim Galore and cutadapt for adapter and quality trimming, and PRINSEQ for sample demultiplexing. In addition, Bowtie2 v2.2.1 (55) was used to map reads to custom databases for bacteria, viruses, human, and vectors. Reads whose highest identity match was not bacterial were removed from subsequent analysis. For bacterial reads, the highest identity match was chosen. If there were multiple top hits, the lowest common ancestor was determined.

### Plasma inflammatory markers

Inflammation-related protein biomarkers were determined after unblinding of participants. Measurements were performed by proximity extension assay using the Olink inflammation panel (Olink, Uppsala, Sweden) following manufacturer’s instructions.

### Statistics

#### Sample size

The primary endpoint of the study was immune efficacy. The null hypothesis was that the probability of developing an antibody and/or T cell response to insulin in the oral insulin group equals the probability of developing an antibody and/or T cell response in the placebo group. Based on results of the Pre-POInT study (24), response rates of 20% in the placebo group and 67% in the oral insulin group were assumed. Accordingly, enrolment and randomization of 44 children 1:1 to two treatment groups was expected to be sufficient to reject the null hypothesis with a two-sided Fisher’s exact test at a significance level of 0.05 and a power of 80% and 10% dropout. The sample size estimation was performed using PS Power and Sample Size Calculations software version 3.0.43 (56).

#### Statistical comparisons

To compare continuous variables between the two independent groups, we used the Mann–Whitney *U* test with normal approximation. The Kruskal-Wallis test was applied when the number of groups was larger than two. Differences in categorical variables between the groups were assessed using Fisher’s exact test. Unless otherwise indicated, continuous variables are reported as the median and IQR. The probabilities of time-to-event variables were estimated with the Kaplan–Meier method and compared with the log-rank test. Additional analyses were planned to compare the immunological outcomes in children with the *INS* AA genotype and treatment effects on the stool microbiome. These and all other analyses were considered exploratory. An interaction between *INS* genotype and treatment on immunological responses to insulin was assessed using the Cox proportional hazards model. All analyses comparing responses in relation to monocyte CD169 expression, and analyses of cell frequency and plasma inflammatory markers were defined post-hoc. Spearman’s correlation was calculated to assess the correlation between two continuous variables. Differences between groups’ centroids defined by a principal component analysis (PCA) were assessed using a permutational multivariate analysis of variance (PERMANOVA). Analysis of age relationships to cell population frequencies included a linear mixed model with the cell frequencies as fixed effects and the children identification numbers as a random effect was fitted to predict the age. Stool analyses were conducted to characterize differences in the microbiome between the two treatment groups, including stratification by *INS* AA genotype and to determine the relationship of the microbiome to the immune responses in blood. Differences in beta diversity were visualized using a principal coordinates analysis (PCoA) followed by a PERMANOVA to assess differences between groups. The significance level of two-sided *P* values was 0.05 for all statistical tests. Point estimates are given together with the 95% CIs.

### Study approval

The study was approved by the Ethikkommission der Fakultät für Medizin der Technischen Universität München (206/15). The parents or legal guardians of each child provided written informed consent prior to inclusion in the study. The study was performed in compliance with the current version of the Helsinki declaration.

## Data Availability

All reasonable requests for raw and analyzed data and materials will be promptly reviewed by the corresponding author to determine whether the request is subject to confidentiality obligations. Any data and materials that can be shared will be available from the corresponding author on reasonable request, with appropriate additional ethical approvals, and released via a material transfer agreement.

## Author contributions

R.A. was the clinical coordinator of the study. J.K., K.L.H., A.E., M.W., C.M., J.R., Y.F., M.H., S.M.H. were responsible for laboratory measurements. A.H. was the medical doctor and M.B. lead study nurse responsible for the clinical trial visits and contacting the families. M.P., K.H., and J.H. were the trial statisticians. A.W. supported the statistical analysis. K.L.H. and J.Z.G. analyzed the microbiome data. J.F.P. oversaw the microbiome analysis. P.A. oversaw the antibody analysis. E.B. oversaw the T cell analysis. A.G.Z. and E.B. were responsible for study design. A.G.Z. and P.A. were responsible for study conduct. E.B. and A.G.Z. were responsible for data analysis. A.G.Z. was the principal investigator of the study. A.G.Z., E.B., and R.A. drafted the manuscript. J.K., K.L.H., J.Z.G., A.E., J.F.P., and P.A. were involved in the interpretation of the results and preparation of the manuscript. All authors read and approved the final version of the manuscript.

## Acknowledgements

We are grateful to the participating families and children; the members of the Data Safety Monitoring Board (Chantal Mathieu, Mark Peakman, Olga Kordonouri); all members of the Pre-POInT-early study team (Stephanie Zillmer, Christian Sebelefsky, Nadezhda Lagoda, Melanie Heinrich, Nicole Maison, Alevtina Durmashkina, Susanne Hivner, Fiona Fischer, Marina Holzmeier, Anita Gavrisan, Claudia Peplow, Ramona Lickert, Marlon Scholz, Yvonne Kriesen, Melanie Herbst), Annett Lindner for technical assistance in measuring gene expression, and the FACS facility of the Center for Molecular Cell Biology, TU Dresden. We thank Lilly Pharmaceuticals for donating the insulin crystals and InPhaSol, Apotheke des Universitätsklinikums Heidelberg for the production and supply of the investigational products. The study was monitored by Viktoria Janke (Münchner Studienzentrum, Technical University Munich). The study sponsor was the Technical University Munich, represented by the School of Medicine. The Pre-POINT-early clinical trial was supported by the German Center for Diabetes Research (DZD e.V.), the Juvenile Diabetes Research Foundation (JDRF grant 1-SRA-2018-546-S-B), and the Federal Ministry of Education and Research (BMBF, grant FKZ01KX1818).

